# Grey- and white-matter resilience to tau, cognition and sex in Alzheimer’s disease

**DOI:** 10.64898/2026.06.16.26355812

**Authors:** S. Boutin, B. Houzé, C. Bedetti, A. Pichet Binette, S.M. Brambati, the Alzheimer’s Disease Neuroimaging Initiative

**Author notes:** Corresponding Author: Simona Maria Brambati, Centre de Recherche de l’Institut Universitaire de Gériatrie de Montréal – 4545 chemin Queen-Mary, Montréal, Québec, Canada, H3W 1W4. Data used in preparation of this article were obtained from the Alzheimer’s Disease Neuroimaging Initiative (ADNI) database (adni.loni.usc.edu). As such, the investigators within the ADNI contributed to the design and implementation of ADNI and/or provided data but did not participate in analysis or writing of this report. A complete listing of ADNI investigators can be found at: http://adni.loni.usc.edu/wp-content/uploads/how_to_apply/ADNI_Acknowledgement_List.pdf.

## Abstract

**INTRODUCTION:** Brain resilience to tau has been mainly studied in relation to grey matter, while its role in white matter remains unclear in Alzheimer’s disease (AD). Sex may moderate associations between brain resilience and cognition.

**METHODS:** We analyzed medial temporal lobe tau PET SUVR, entorhinal cortical thickness, cingulum-hippocampal mean diffusivity, and cognition in 205 amyloid-positive individuals from ADNI. Associations between grey- and white-matter resilience to tau and cognitive performance or decline were examined using linear and mixed-effects models, including sex interactions and stratified analyses.

**RESULTS:** Higher grey-matter resilience to tau related to better cross-sectional memory and language performance (p<0.005), whereas higher white-matter resilience related to slower decline in these domains (p<0.03). These associations were seen in men but not women. DISCUSSION: White-matter resilience may better capture early cognitive changes in AD. Sex differences suggest greater sensitivity of cognition to brain resilience in men.

## 1. Background

Alzheimer’s disease (AD) is characterized by the progressive accumulation of beta-amyloid (Aβ) plaques and tau tangles, which start decades before symptom onset [1]. In preclinical and prodromal stages, cognitively unimpaired individuals (CU) or those with mild cognitive impairment (MCI) who exhibit higher Aβ levels are at increased risk of AD dementia [1–4]. According to the amyloid cascade hypothesis [5,6], Aβ accumulation initiates tau aggregation, which in turn leads to neurodegeneration and eventually cognitive decline. According to Braak stages [7], tau pathology initially accumulates in medial temporal areas, and previous studies have shown that neurodegeneration also happen in those same regions (e.g., entorhinal cortex, hippocampus) [8–10]. Beyond the effect of tau on atrophy, previous evidence has also shown a link between higher tau levels and white-matter-microstructure degeneration in early AD [11,12], particularly increases in mean diffusivity (MD) of the tracts [13]. White-matter microstructural changes might even precede tau accumulation or lead to tau propagation from medial temporal regions to the neocortex [12].

The timing of these pathological events – and the resulting cognitive impairment – varies across individuals. For example, the relationship between tau accumulation and brain atrophy may differ from person to person, with some individuals showing greater resilience to pathological burden. Understanding this variability is essential for clarifying how these processes translate into the clinical manifestations of AD. Brain resilience is defined as better-than-expected brain integrity given pathological burden [14]. [8–10]Cortical thickness and mean diffusivity of white matter tracts of medial temporal regions can thus be useful indicators of brain resilience to the earliest sites of tau deposition [15]. However, most studies investigating brain resilience were not focused on brain resilience to tau. Higher grey-matter resilience (e.g., greater whole-brain cortical thickness or hippocampal volume) to tau has been linked to better cognitive outcomes and lower risk of clinical progression [16–18]. Comparatively, no studies have deeply investigated white-matter resilience to tau and its link with cognitive and clinical outcomes. Only one study reported that greater white-matter resilience (i.e., more intact macro- and microstructural white matter in limbic and default mode networks) attenuates the association between tau burden and memory decline [19]. As AD can be conceptualized as a network-based disease [20–22], it is also important to examine white-matter resilience and not only grey matter. [20–22]Most studies on brain resilience also focused on global cognition or memory, leaving the role of brain resilience in other cognitive domains (e.g., language, executive function) less clear. Although memory is typically affected earliest in AD, impairments in other domains, particularly language, also emerge early and are associated with pathology [23,24]. Moreover, [20–22]individual variables such as education level, age and APOE-ε4 status have been shown to contribute to brain resilience variability [17,25]. However, sex is relatively less studied compared to other individual factors. Evidence is mixed regarding sex differences in grey-matter resilience: some studies report no differences [25], whereas others suggest women [17] or men exhibit higher resilience [26]. This variability may reflect the disease stage examined, as women often show preserved brain structure in early stages or at low pathology levels but appear to lose this advantage as pathology increases [27–32]. Thus, sex-specific analyses at preclinical and prodromal AD stages are necessary to clarify how sex influences the relationship between tau and neurodegeneration. Understanding sex differences in brain resilience may provide insights at the anatomical level into sex-specific cognitive trajectories along the AD continuum.

Here, we investigate grey- and white-matter resilience to tau in relation to cognitive performance and cognitive decline over time across several domains (memory, language, executive function and visuospatial abilities), as well as whether there are sex differences in those associations.

## 2. Methods

Data used in the preparation of this article were obtained from the Alzheimer’s Disease Neuroimaging Initiative (ADNI) database (adni.loni.usc.edu). The ADNI was launched in 2003 as a public-private partnership, led by Principal Investigator Michael W. Weiner, MD. The primary goal of ADNI has been to test whether serial magnetic resonance imaging (MRI), positron emission tomography (PET), other biological markers, and clinical and neuropsychological assessment can be combined to measure the progression of mild cognitive impairment (MCI) and early Alzheimer’s disease (AD). For up-to-date information, see www.adni-info.org. The data supporting the findings of this study are not publicly available in respect to Alzheimer’s Disease Neuroimaging Initiative data sharing and publication policy. A request made to the ADNI data committee needs to be filled in order to access this data. However, to facilitate openness, transparency and reproducibility of research, the code used to clean and analyze the data supporting the findings of this study are openly available in Open Science Framework at https://osf.io/3sfzb/overview?view_only=6d8d73352c2d44a4bf0d704df622539b

### 2.1 Participants

We included CU and MCI individuals who were categorized as Aβ positive (Aβ-PET Standardized Uptake Value Ratio [SUVR] greater than 1.11 in a cortical summary region that includes frontal, anterior/posterior cingulate, lateral parietal and lateral temporal regions) from the ADNI database. In the general ADNI inclusion criteria, participants are considered CU if they present: 1) a Mini-Mental State Examination score between 24 and 30; 2) a Clinical Dementia Rating score of 0; 3) above education-adjusted scores on the delayed Paragraph Recall task from the Wechsler Memory Scale Logical Memory II and 4) no significant impairment in cognitive functions or activities of daily living. To be included in the MCI group, participants must present: 1) memory complaints that are verified by their study partner; 2) a Mini-Mental State Examination score between 24 and 30; 3) a Clinical Dementia Rating score of 0.5; 4) below education-adjusted scores on the delayed Paragraph Recall task from the Wechsler Memory Scale Logical Memory II and 5) sufficiently preserved cognitive functions and functional performance such that AD diagnosis cannot be made. Study-specific inclusion criteria include having available tau-PET, structural magnetic resonance imaging (grey matter), diffusion imaging (white matter microstructure) and cognitive data. The ADNI protocols were approved by all the Institutional Review Boards of the participating institutions. All participants provided written informed consent prior to enrolment in the study. For full description of Institutional Review Boards procedures and approvals, see: https://adni.loni.usc.edu/help-faqs/adni-documentation/. In addition to ADNI ethical considerations, this study was also approved by the Education and Psychology Research Ethics Committee of *Université de Montréal* (CEREP) and by the Aging-Neuroimaging Research Ethics Committee of the Centre Intégré Universitaire de Santé et de Services Sociaux du Centre-Sud-de-l’Île-de-Montréal.

### 2.2 Tau PET

To quantify tau burden, we used the mean SUVR of both the entorhinal cortex and amygdala from PET, as these regions are among the first affected by tau pathology [33]. We used PET data available with the flortaucipir tracer and processed from the ADNI PET core. The inferior cerebellar grey matter was used as the reference region to generate SUVR. The complete descriptions of the collection and analyses protocols are provided in the ADNI procedural manual at www.adni-info.org.

Since all imaging exams and cognitive assessments were not done at the same time point, we kept only MRI and cognitive measures that were available within a year (before or after) of the first tau-PET scan.

### 2.3 MRI measures

#### 2.3.1 Grey matter

To assess grey matter atrophy, we used the cortical thickness of the entorhinal cortex (mean of left and right cortices) obtained with structural MRI [34]. We used available MRI data in ADNI which was processed with FreeSurfer version 7.2.0 (https://surfer.nmr.mgh.harvard.edu/fswiki/rel7downloads). Cortical thickness appears to be a more reliable measure of atrophy than volume, as the relatively low variability in grey matter cytoarchitecture allows it to more directly reflect pathological changes and neurodegeneration [35–37]. We chose the entorhinal cortex as our region of interest to be consistent with the regions selected for tau measurement.

#### 2.3.2 White matter

We used available pre-processed diffusion-weighted MRI data (MD) released by ADNI team at University of Southern California. MD maps were estimated from corrected data with FSL’s dtifit. White matter region-of-interest measures were derived from the JHU ICBM-DTI-81 atlas, which was non-linearly registered to each participant’s diffusion space, and robust means were computed for each region of interest [38]. Here, we specifically used the MD measure of the cingulum-hippocampal connections (mean left and right) to assess white matter microstructure. We chose the cingulum-hippocampal connections as our tract of interest, as it is the one most closely related to the tau region of interest.

#### 2.3.3 Resilience measures

To obtain a measure of grey- and white-matter resilience to tau, we extracted the residuals of the associations between tau levels and 1) mean entorhinal cortical thickness and 2) MD of the cingulum-hippocampal connections, using two separate linear regression models on the whole sample, controlling for age (Supplemental Figure 1). Conceptually, the residuals represent higher- or lower-than-expected cortical thickness, as well as lower- or higher-than-expected white-matter diffusivity based on tau burden, reflecting high or low brain resilience, respectively. We compared residuals from the full sample with those derived from sex-specific models (i.e., male and female subsample only), and both approaches were strongly correlated (r>0.98; Supplemental Figures 2 and 3). Accordingly, residuals from the full-sample models were used to ensure greater consistency across analyses.

**Figure 1.**
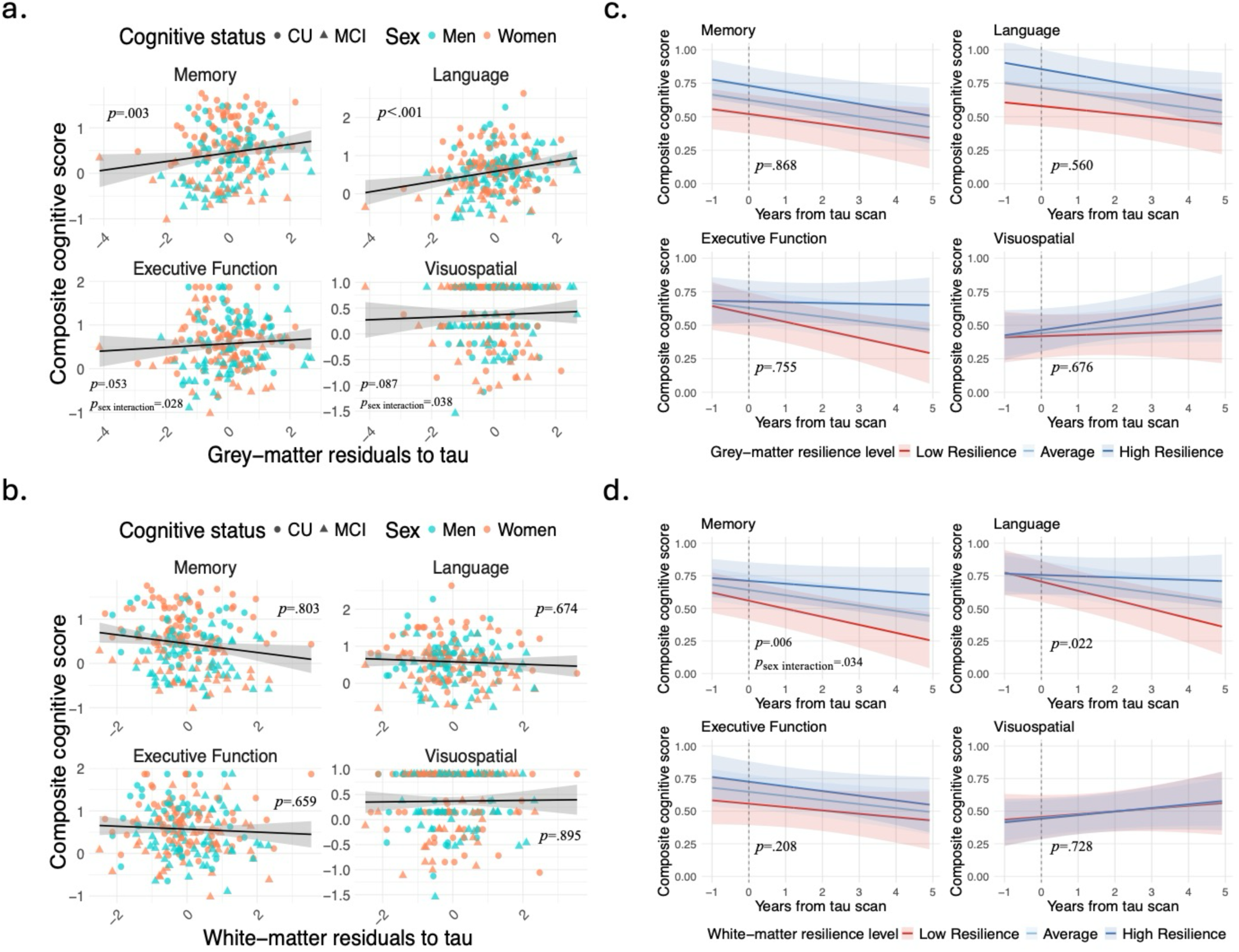
Associations between grey- and white-matter resilience to tau and cognitive performance and decline over time across domains. Note. Reported *p* values reflect the associations between grey- (a) and white-matter (b) resilience to tau and cognitive performance, as well as between grey- (c) and white-matter (d) resilience and cognitive decline over time. For the sex interaction, only significant *p* values are shown. In panels c and d, the grey dashed line represents the timepoint of the tau scan. For visualization purposes only, we created a brain resilience level measure by splitting the obtained grey- and white-matter residuals into tertiles, where individuals in the bottom tertile were categorized as having “low resilience” and those in the top tertile were considered as having “high resilience”. To facilitate interpretation, we flipped the sign of the white-matter residuals (i.e., low residuals [lower mean diffusivity of the cingulum-hippocampal connections than expected based on tau levels] became high resilience).

**Figure 2.**
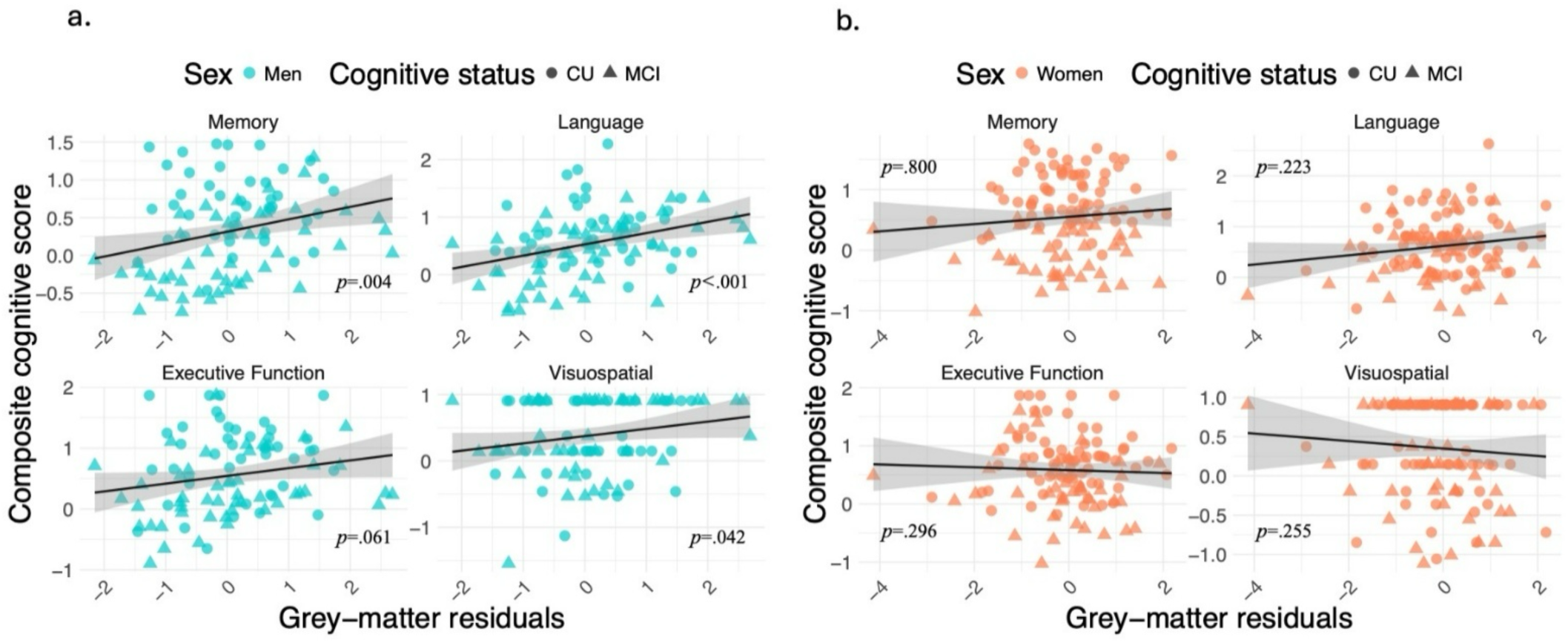
Associations between grey-matter resilience and cognitive performance across domains separately in men (a) and women (b).

**Figure 3.**
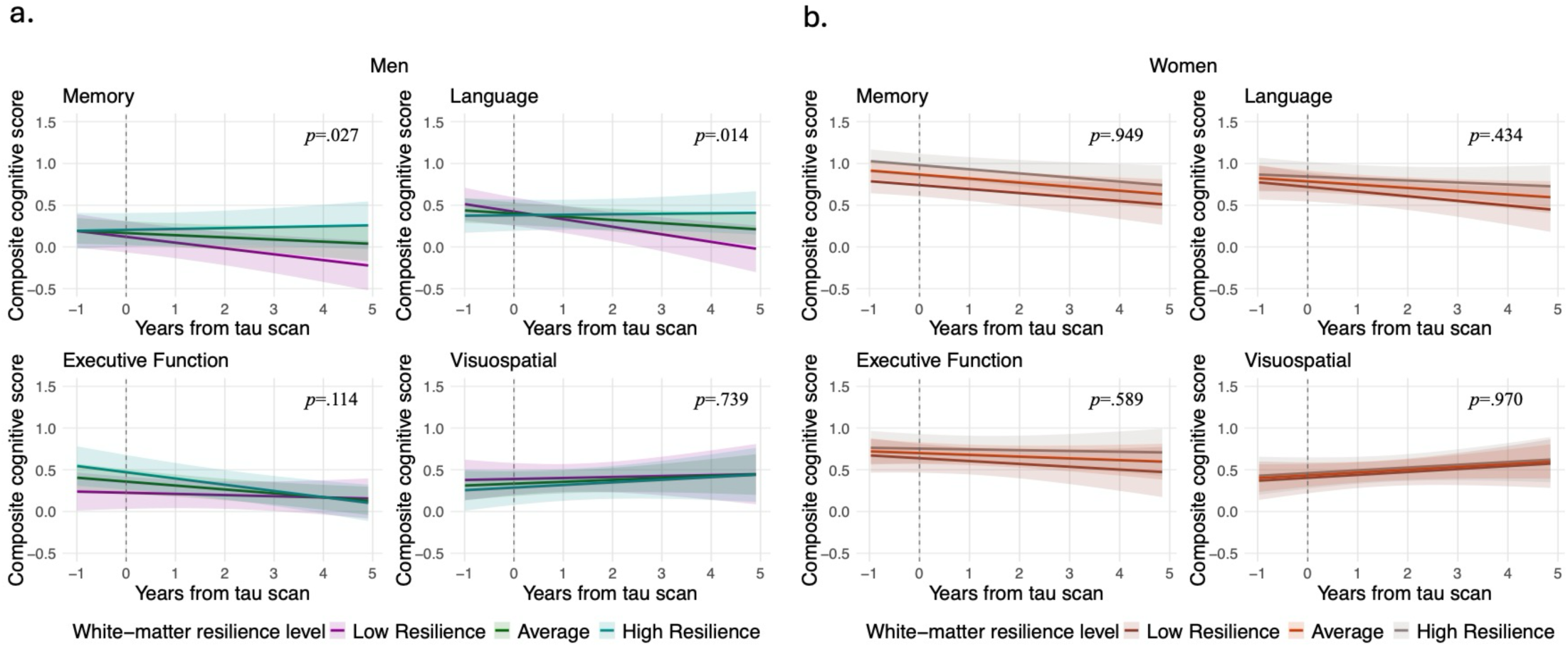
Associations between white-matter resilience and cognitive decline over time across domains separately in men (a) and women (b). Note. The grey dashed line represents the timepoint of the tau scan. For visualization purposes only, we created a brain resilience level measure by splitting the obtained grey-and white-matter residuals into tertiles, where individuals in the bottom tertile were categorized as having “low resilience” and those in the top tertile were considered as having “high resilience”. To facilitate interpretation, we flipped the sign of the white-matter residuals (i.e., low residuals [lower mean diffusivity of the cingulum-hippocampal connections than expected based on tau levels] became high resilience).

### 2.4 Cognition

To assess cognitive performance in multiple domains, we used the derived and validated composite scores available in ADNI, and we selected the cognitive timepoint closest to the first available tau scan. To assess cognitive decline over time, we included participants who had at least 2 cognitive assessments that were performed up to 1 year before and 5 years after the tau scan, thereby maximizing the number of participants with longitudinal follow-up data. To obtain composite cognitive scores in ADNI, anchored items and neuropsychological tests were categorized into four cognitive domains: memory, language, executive function, and visuospatial abilities, which allows summarizing performance. For a complete description on how these scores were calculated, please refer to [39].

It is important to note that the visuospatial composite score exhibited an atypical distribution, with markedly many individuals capped at the highest value of 1 or having the same score cross-sectionally and over time compared to the composite scores in the other cognitive domains. This irregularity was also noticed by previous authors [40], and likely reflects characteristics of the items used in the calculation of this composite score, as 5 out of 6 items were yes or no questions, limiting the variability. Although we report all analyses involving this measure for transparency, results related to the visuospatial domain should be interpreted cautiously, as the distribution may introduce bias into subsequent analyses.

### 2.5 Statistical analyses

All statistical analyses were performed using R version 4.2.2. Two-sample t-tests or chi-squared tests, accordingly, were performed to determine whether there were sex differences on demographics and variables of interest, both in cross-sectional analyses and the longitudinal subsample (at baseline).

To examine the associations between grey- and white-matter resilience to tau and cognitive performance across multiple domains (memory, language, executive function, and visuospatial abilities), we first performed whole-group cross-sectional analyses using linear regression models with the derived residuals. All models included an interaction term between derived residuals and sex, as well as age, years of education and cognitive status (CU or MCI) as covariates. Separate models were used for grey- and white-matter resilience measures and for each of the four cognitive domains. Then, using a subsample with available data, we conducted longitudinal analyses to examine the association between grey- and white-matter resilience and cognitive decline over time using linear mixed-effects models with random intercepts and slopes. Such models included an interaction term of residuals, sex and time, along with age at baseline, years of education, and cognitive status. Here, time is defined as the interval in years between the tau scan and each cognitive assessment.

To further examine the effect of sex, we complemented with sex-stratified analyses, conducting all cross-sectional and longitudinal models separately in men and women.

Two-sided p-value < 0.05 were considered significant.

## 3. Results

### 3.1 Description of participants and characterization of brain resilience

In total, 93 men (43 CU and 50 MCI) and 112 women (70 CU and 42 MCI) who were Aβ positive were included in the present study. Longitudinal cognitive data was available in a subsample of 68 men (30 CU and 38 MCI) and 83 women (52 CU and 31 MCI) (Table 1). The two-sample t-tests and chi-squared tests showed that there were more CU individuals in women [χ^2^(1)=4.80, *p=*.029], that men were older [t(198.88)=3.28, *p=*.001], and that women had higher memory composite scores [t(201.38)=-2.54, *p=*.012]. Sex differences in the longitudinal subsample were comparable (Table 1).

**Table 1.**
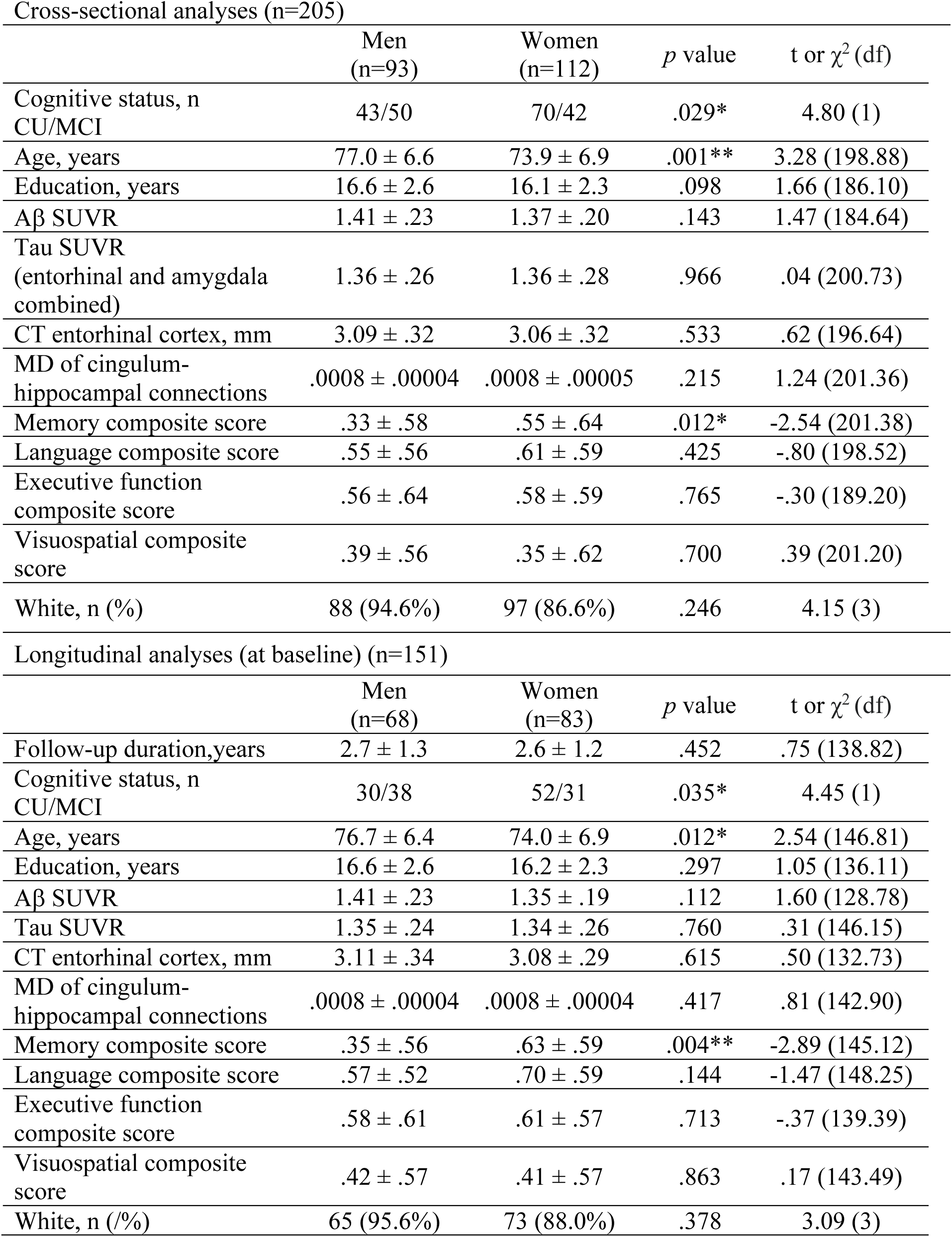
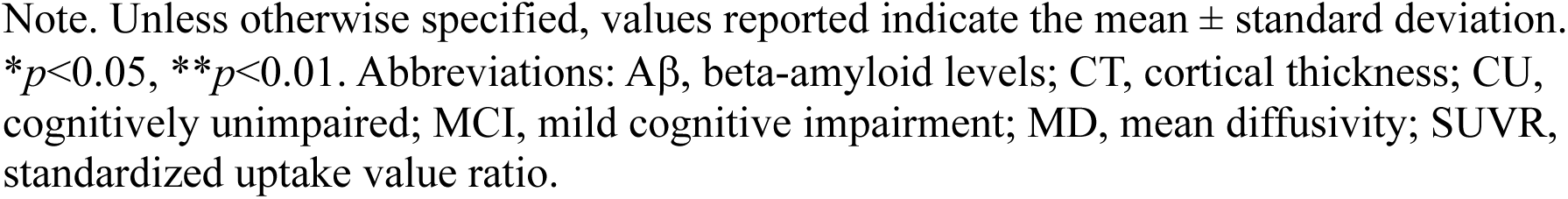
Subject demographics and characteristics for cross-sectional and longitudinal analyses (at baseline), split by sex. Note. Unless otherwise specified, values reported indicate the mean ± standard deviation. \**p*<0.05, \*\**p*<0.01. Abbreviations: Aβ, beta-amyloid levels; CT, cortical thickness; CU, cognitively unimpaired; MCI, mild cognitive impairment; MD, mean diffusivity; SUVR, standardized uptake value ratio.

| Cross-sectional analyses (n=205) |  |  |  |  |
| --- | --- | --- | --- | --- |
| | Men<br>(n=93) | Women<br>(n=112) | *p* value | t or $\chi^2$ (df) |
| Cognitive status, n<br>CU/MCI | 43/50 | 70/42 | .029* | 4.80 (1) |
| Age, years | 77.0 ± 6.6 | 73.9 ± 6.9 | .001** | 3.28 (198.88) |
| Education, years | 16.6 ± 2.6 | 16.1 ± 2.3 | .098 | 1.66 (186.10) |
| Aβ SUVR | 1.41 ± .23 | 1.37 ± .20 | .143 | 1.47 (184.64) |
| Tau SUVR<br>(entorhinal and amygdala<br>combined) | 1.36 ± .26 | 1.36 ± .28 | .966 | .04 (200.73) |
| CT entorhinal cortex, mm | 3.09 ± .32 | 3.06 ± .32 | .533 | .62 (196.64) |
| MD of cingulum-hippocampal connections | .0008 ± .00004 | .0008 ± .00005 | .215 | 1.24 (201.36) |
| Memory composite score | .33 ± .58 | .55 ± .64 | .012* | -2.54 (201.38) |
| Language composite score | .55 ± .56 | .61 ± .59 | .425 | -.80 (198.52) |
| Executive function<br>composite score | .56 ± .64 | .58 ± .59 | .765 | -.30 (189.20) |
| Visuospatial composite<br>score | .39 ± .56 | .35 ± .62 | .700 | .39 (201.20) |
| White, n (%) | 88 (94.6%) | 97 (86.6%) | .246 | 4.15 (3) |
| Longitudinal analyses (at baseline) (n=151) |  |  |  |  |
| | Men<br>(n=68) | Women<br>(n=83) | *p* value | t or $\chi^2$ (df) |
| Follow-up duration, years | 2.7 ± 1.3 | 2.6 ± 1.2 | .452 | .75 (138.82) |
| Cognitive status, n<br>CU/MCI | 30/38 | 52/31 | .035* | 4.45 (1) |
| Age, years | 76.7 ± 6.4 | 74.0 ± 6.9 | .012* | 2.54 (146.81) |
| Education, years | 16.6 ± 2.6 | 16.2 ± 2.3 | .297 | 1.05 (136.11) |
| Aβ SUVR | 1.41 ± .23 | 1.35 ± .19 | .112 | 1.60 (128.78) |
| Tau SUVR | 1.35 ± .24 | 1.34 ± .26 | .760 | .31 (146.15) |
| CT entorhinal cortex, mm | 3.11 ± .34 | 3.08 ± .29 | .615 | .50 (132.73) |
| MD of cingulum-hippocampal connections | .0008 ± .00004 | .0008 ± .00004 | .417 | .81 (142.90) |
| Memory composite score | .35 ± .56 | .63 ± .59 | .004** | -2.89 (145.12) |
| Language composite score | .57 ± .52 | .70 ± .59 | .144 | -1.47 (148.25) |
| Executive function<br>composite score | .58 ± .61 | .61 ± .57 | .713 | -.37 (139.39) |
| Visuospatial composite<br>score | .42 ± .57 | .41 ± .57 | .863 | .17 (143.49) |
| White, n (/%) | 65 (95.6%) | 73 (88.0%) | .378 | 3.09 (3) |

Note. Unless otherwise specified, values reported indicate the mean ± standard deviation. \* $p < 0.05$ , \*\* $p < 0.01$ . Abbreviations: Aβ , beta-amyloid levels; CT, cortical thickness; CU, cognitively unimpaired; MCI, mild cognitive impairment; MD, mean diffusivity; SUVR, standardized uptake value ratio.

Higher tau PET levels were associated with both lower entorhinal cortical thickness [β(SE)=-.375(.080), *p*<.001, β_standardized_=-.317] and higher mean diffusivity (MD) of cingulum-hippocampal connections [β(SE)=.00005(.00001), *p*<.001, β_standardized_=.272; see Supplemental Figure 1]. Grey- and white-matter resilience to tau were not correlated, as there was no significant association between grey- and white-matter residuals (*r*(203)=-.08, *p=*.273; see Supplemental Figure 4). We also found no sex differences in grey- [t(197.22)=1.43, *p=*.155] nor white-matter resilience [t(201.79)=.15, *p=*0.882] (Supplemental Figure 5).

### 3.2 Resilience to tau in relation to cognitive performance and the effect of sex

#### 3.2.1 Grey-matter resilience

Higher grey-matter resilience to tau (i.e., greater-than-expected entorhinal cortical thickness given tau levels) was associated with higher cognitive performance in memory [β(SE)=.142(.047), *p=*.003, β_standardized_=.230, R^2^_adjusted=_.488] and language domains [β(SE)=.186(.054), *p<*.001, β_standardized_=.324, R^2^_adjusted=_.216] (Figure 1a; Supplemental Table 1).

Looking at sex differences, there were significant interactions suggesting a stronger protective effect of grey-matter resilience to tau on executive functioning [β(SE)=-.172(.078), *p=*.028, β_standardized_=-.210] and visuospatial abilities [β(SE)=-.173(.083), *p=*.038, β_standardized_=-.218] in men than in women (Figure 1a; Supplemental Table 1). In sex-stratified analyses, higher grey-matter resilience was associated with higher memory [β(SE)=.144(.049), *p=*.004, β_standardized_=.247], language [β(SE)=.190(.051), *p<*.001, β_standardized_=.333] and visuospatial [β(SE)=.120(.058), *p=*.042, β_standardized_=.212] performance among men (Figure 2a; Supplemental Table 2). On the other hand, grey-matter resilience to tau was not associated with cognitive performance in any domains among women (Figure 2b; Supplemental Table 2).

#### 3.2.2 White-matter resilience

White-matter resilience to tau (i.e., lower-than-expected MD of cingulum-hippocampal connections given tau levels) was not associated with cognitive performance in any domain (Figure 1b; Supplemental Table 3).

Regarding the effect of sex, no interactions were found between sex and white-matter residuals in any of the cognitive domains (Supplemental Table 3). In sex-stratified analyses, white-matter resilience to tau was not associated with cognitive performance in any domain among men, but higher white-matter resilience was associated with higher memory performance among women [β(SE)=-.086(.037), *p=*.022, β_standardized_=-.142] (Supplemental Figure 6; Supplemental Table 4).

### 3.3 Resilience to tau in relation to cognitive decline over time and the effect of sex

#### 3.3.1 Grey-matter resilience

On a subsample of participants with available longitudinal cognitive data (74% of the whole sample), there was no association between grey-matter resilience to tau and cognitive decline over time in any domain (Figure 1c; Supplemental Table 5), and this longitudinal association did not vary by sex (Supplemental Table 5). The results of the linear mixed-effects models also mostly aligned with those of the cross-sectional analyses regarding the effect of grey-matter resilience and sex differences at baseline, while additionally indicating that women exhibited better memory performance (see Supplemental Table 5).

In sex-stratified analyses, there was also no association between grey-matter resilience to tau and cognitive decline in any domain among men (Supplemental Figure 7; Supplemental Table 6). Among women, higher grey-matter resilience to tau was associated with slower cognitive decline in executive functioning [β(SE)=.048 (.022), *p*=.034, β_standardized_=.11] (Supplemental Figure 7; Supplemental Table 6).

#### 3.3.2 White-matter resilience

We found that higher white-matter resilience to tau was associated with slower cognitive decline over time in memory [β(SE)=-.043 (.015), *p*=.006, β_standardized_=-.09] and language [β(SE)=-.045 (.019), *p*=.022, β_standardized_=-.11] (Figure 1d; Supplemental Table 8). The results also showed that the association between white-matter resilience and memory decline varied by sex [β(SE)=.044 (.020), *p*=.034] (Figure 1d; Supplemental Table 7), where the effect of higher white-matter resilience on slowing cognitive decline was greater in men than women. Otherwise, the results mostly aligned with those of the cross-sectional analyses regarding the effect of white-matter resilience and sex differences at baseline, while again additionally indicating that women exhibited better memory performance (see Supplemental Table 7).

In sex-stratified analyses (Figure 3; Supplemental Table 8), there was an association between white-matter resilience to tau and slower cognitive decline in memory [β(SE)=-.038 (.017), *p*=.027, β_standardized_=-.08] and language [β(SE)=-.045 (.018), *p*=.014, β_standardized_=-.11] among men, while there were no associations in women.

### 3.4 Post-hoc analyses

Based on our findings linking cognitive performance and decline to grey- and white-matter resilience, respectively, and given that resilience was operationalized as the residuals from the associations between tau burden and 1) cortical thickness and 2) white-matter MD, we further examined whether these associations were simply driven by the well-established relationship between higher tau levels and worse cognitive outcomes [41,42]. To do so, we first performed linear regressions by including both tau and entorhinal cortical thickness as predictors of cognitive performance. These results showed that, although lower tau was indeed associated with higher cognitive performance in almost all domains [memory; β(SE)=-.735(.124), *p*<.001, β_standardized_=-.318, executive function; β(SE)=-.408(.167), *p*=.015, β_standardized_=-.179, visuospatial; β(SE)=-.359(.178), *p*=.045, β_standardized_=-.163], greater entorhinal cortical thickness was also associated with better memory [β(SE)=.266(.095), *p*=.006, β_standardized_=.136] and language performance [β(SE)=.399(.120), *p*=.001, β_standardized_=.220], above and beyond the effect of tau.

Similarly, we examined whether our white-matter variable of interest was associated with cognitive decline over time, above and beyond tau burden. We performed linear mixed-effects models including interactions between time and tau, as well as time and MD of cingulum-hippocampal connections predicting domain-specific cognitive decline. These results showed that, although lower tau was indeed associated with slower decline in the memory domain [β(SE)=-.211(.047), *p*<.001, β_standardized_=-.11], lower MD of cingulum-hippocampal connections was associated with slower language decline [β(SE)=-643.9(289.2), *p*=.028, β_standardized_=-.07], above and beyond the effect of tau on cognitive decline.

## 4. Discussion

In this study, we examined grey- and the less studied white-matter resilience to tau in relation to cognitive performance and decline over time across several domains, as well as the effect of sex on these associations, in a sample of CU and MCI Aβ positive individuals. With regards to cognitive performance, greater grey-matter resilience to tau, but not white-matter, was associated with better memory and language. Our findings revealed a reverse pattern for cognitive decline over time: greater white-matter resilience, but not grey-matter, was associated with slower decline in both memory and language. With respect to sex, although there were no sex differences in the level of brain resilience to tau, its relationship, particularly in relation to grey matter, with cognitive outcomes varied between sexes. Grey-matter resilience to tau was more strongly associated with better cognitive performance and slower decline across different domains in men than in women. Together, these findings highlight the importance of incorporating measures of white-matter resilience, particularly for predicting cognitive trajectories over time, alongside the traditionally studied grey-matter, to better capture heterogeneity in disease expression and clinical trajectories at the early AD stages. They also underscore the need to consider sex as a key factor interacting with pathology burden and resilience to shape cognitive outcomes.

Consistent with prior work, our findings replicate the association between greater grey-matter resilience to tau and better cognitive outcomes [16–18]. However, a novel aspect of the present study is our focus on cortical thickness resilience in the entorhinal cortex, a key medial temporal region, rather than on global cortical atrophy. This region-specific approach is particularly relevant given that tau pathology accumulates early and preferentially in medial temporal structures in AD, allowing for a more precise characterization of brain resilience to tau by targeting areas most vulnerable to early neurodegeneration [8–10]. Our results also extend previous findings by showing that grey-matter resilience related to cognitive performance beyond the memory domain, with language abilities emerging as a particularly relevant outcome. The novelty of our results thus lies in the inclusion of cognitive composite scores across multiple domains, rather than focusing solely on memory, but also stems from the use of a more precise measure of grey-matter resilience in a brain region particularly affected in early AD. Prior studies have shown that subtle language changes, including alterations in connected speech, naming, and semantic processing, can be detected as early AD manifestations [23,24,43,44], hence the importance of studying this domain. Our results support an association between brain resilience to tau and preserved language function, further emphasizing the link between language impairments and tau-induced brain pathology.

Importantly, we extend resilience models by incorporating a measure of white matter, a comparatively underexplored domain, thereby providing a more comprehensive framework for understanding the neural substrates of cognitive outcomes. Although white-matter resilience to tau was not associated with cross-sectional cognitive performance, we found that greater resilience was associated with slower decline over time in both memory and language functions. To our knowledge, only one prior study has similarly reported that higher white-matter resilience attenuates the association between tau pathology and memory decline over time [19]. Their findings provided preliminary evidence of a link between white-matter resilience and cognitive trajectories in CU individuals. Here, we provide clearer evidence by again showing that language trajectories are also sensitive to brain resilience and extend this work by including individuals with MCI, who are further along the AD continuum. The lack of association between grey-matter resilience and cognitive decline, coupled with the specific longitudinal associations observed for white-matter resilience, may suggest that microstructural white-matter alterations (e.g., higher MD) represent earlier or more sensitive markers of disease progression, potentially linked to the spread of pathology through brain networks. While current theoretical models of AD describe the sequence of pathological processes underlying disease progression, our study extends this framework by examining how grey- and white-matter resilience may help better understand heterogeneity in this timing of events. Tau-related neuronal damage may contribute to cognitive impairments at a given timepoint, whereas white-matter degeneration may be more closely linked to cognitive changes over time [45]. Growing evidence supports the conceptualization of AD as a network-based disease [20–22], underscoring the importance of examining how tau pathology disrupts structural connectivity and, in turn, shapes longitudinal cognitive trajectories.

Another contribution of this study to AD models is the demonstration of the importance of sex in moderating the associations between brain resilience and cognitive outcomes. Specifically, our findings suggest that grey-matter resilience may play a more prominent role in preserving cognitive functions in men than in women. A prior preliminary study reported sex-specific predictors of resistance to tau propagation, with cognition, education, and APOE-ε4 status being more influential in women, and age and Aβ burden being more influential in men [15]. Building on this work, we extend the literature by moving beyond the identification of sex differences in resilience per se to examine how sex shapes cognitive profiles and trajectories as a function of brain resilience. The literature on sex differences in white-matter resilience remains particularly limited. Some prior evidence indicates sex differences in microstructural changes in AD [46,47], yet studies linking AD pathology and white matter integrity typically adjust for sex rather than testing sex-specific effects [11,12,48]. In the present study, sex influenced the associations between grey- and white-matter resilience and cognitive performance and decline, respectively, highlighting the importance of incorporating both interaction terms and sex-stratified approaches in future studies to more fully characterize sex-specific effects. Although our findings and prior work suggest that the pathological mechanisms linking tau accumulation to grey- and white-matter degeneration are broadly similar between men and women, with no consistent sex differences in brain resilience [17,25,26], the cognitive impact of resilience, particularly in grey matter, on memory and language performance appears to favor men. Thus, while women in our sample demonstrated better memory performance overall, their cognitive advantage did not appear to be strongly modulated by higher levels of brain resilience. Additional factors beyond brain resilience to tau, such as hormonal influences, vascular health, or other sex-specific protective mechanisms, may thus be particularly important for understanding cognitive resilience in women. Indeed, previous studies have identified sex-specific risk and protective factors for cognitive decline that also vary according to cognitive status and pathology burden [49–52]. Along the AD continuum, women typically show a verbal memory advantage at similar pathology levels early in the disease [53–55], but tend to lose this advantage and experience faster decline later on [56–58]. The exact mechanisms underlying this shift remain unclear, although our results on sex-specific associations between brain resilience and cognitive outcomes might help clarify these sex-specific cognitive trajectories at the anatomical level, as a stronger effect of resilience in preserving cognitive functions in men may contribute to their cognitive advantage over women at later pathological and clinical stages [49,59].

Some limitations of the study should be acknowledged. The correlational design precludes causal inferences, and we cannot conclude that greater brain resilience directly causes better cognitive outcomes. Although our main objective was to focus on early AD stages, tau quantity is typically lower among CU and MCI individuals, which may have limited our ability to detect stronger or more widespread associations. Future studies could explore these associations along the whole AD continuum for a more comprehensive understanding. The use of bigger databases with longer follow-up could help better examine whether brain resilience predicts clinical progression (e.g., from CU to MCI, or from MCI to AD). Although cognitive composite scores facilitate domain-level analyses, their clinical utility may be limited, as clinical practice typically relies on individual neuropsychological measures. More representative indices of visuospatial abilities are also needed, as the atypical distribution of these scores, which could stem from an error in the database, limits interpretability. Finally, the ADNI cohort presents limited racial and ethnic diversity, which may restrict the generalizability of our findings to more diverse populations and underscores the need for future studies in more socioeconomically and ethnically diverse cohorts.

In conclusion, this study provides new evidence that brain resilience to tau is a multidimensional construct with distinct and complementary contributions from grey-and white-matter integrity, and that its cognitive relevance extends beyond memory to include other domains, most notably language. By adopting a region-specific approach to grey-matter resilience and incorporating the comparatively understudied white-matter component, our findings demonstrate that grey-matter resilience is primarily associated with cross-sectional cognitive performance, whereas white-matter resilience is more closely related to longitudinal cognitive trajectories. These results support network-based models of AD and highlight the importance of considering both cortical and connective substrates when characterizing resilience to tau. Importantly, our findings further show that sex moderates the cognitive impact of brain resilience, suggesting that similar levels of neuropathology may translate into different cognitive profiles and trajectories in men and women. Together, this work advances current models of resilience by emphasizing the need to integrate white-matter measures and sex-specific approaches to more accurately capture heterogeneity in disease expression and to better predict cognitive outcomes along the AD continuum.

## Acknowledgments/Funding Sources/Disclosures

### a) Acknowledgments

Data collection and sharing for this project was funded by the Alzheimer’s Disease Neuroimaging Initiative (ADNI) (National Institutes of Health Grant U01 AG024904) and DOD ADNI (Department of Defense award number W81XWH-12-2-0012). ADNI is funded by the National Institute on Aging, the National Institute of Biomedical Imaging and Bioengineering, and through generous contributions from the following: AbbVie, Alzheimer’s Association; Alzheimer’s Drug Discovery

Foundation; Araclon Biotech; BioClinica, Inc.; Biogen; Bristol-Myers Squibb Company; CereSpir, Inc.; Cogstate; Eisai Inc.; Elan Pharmaceuticals, Inc.; Eli Lilly and Company; EuroImmun; F. Hoffmann-La Roche Ltd and its affiliated company Genentech, Inc.; Fujirebio; GE Healthcare; IXICO Ltd.; Janssen Alzheimer Immunotherapy Research & Development, LLC.; Johnson & Johnson Pharmaceutical Research & Development LLC.; Lumosity; Lundbeck; Merck & Co., Inc.; Meso Scale Diagnostics, LLC.; NeuroRx Research; Neurotrack Technologies; Novartis Pharmaceuticals Corporation; Pfizer Inc.; Piramal Imaging; Servier; Takeda

Pharmaceutical Company; and Transition Therapeutics. The Canadian Institutes of Health Research is providing funds to support ADNI clinical sites in Canada. Private sector contributions are facilitated by the Foundation for the National Institutes of Health (www.fnih.org). The grantee organization is the Northern California Institute for Research and Education, and the study is coordinated by the Alzheimer’s Therapeutic Research Institute at the University of Southern California. ADNI data are disseminated by the Laboratory for Neuro Imaging at the University of Southern California.

## Data Availability

The data supporting the findings of this study are not publicly available in respect to Alzheimer's Disease Neuroimaging Initiative data sharing and publication policy. A request made to the ADNI data committee needs to be filled in order to access this data. However, to facilitate openness, transparency and reproducibility of research, the code used to clean and analyze the data supporting the findings of this study are openly available in Open Science Framework.

https://osf.io/3sfzb/overview?view_only=6d8d73352c2d44a4bf0d704df622539b

