## Supplementary Material for "Grey- and white-matter resilience to tau, cognition and sex in Alzheimer’s disease"

Supplemental Figure 1. Associations between tau levels and grey- and white-matter variables of interest.

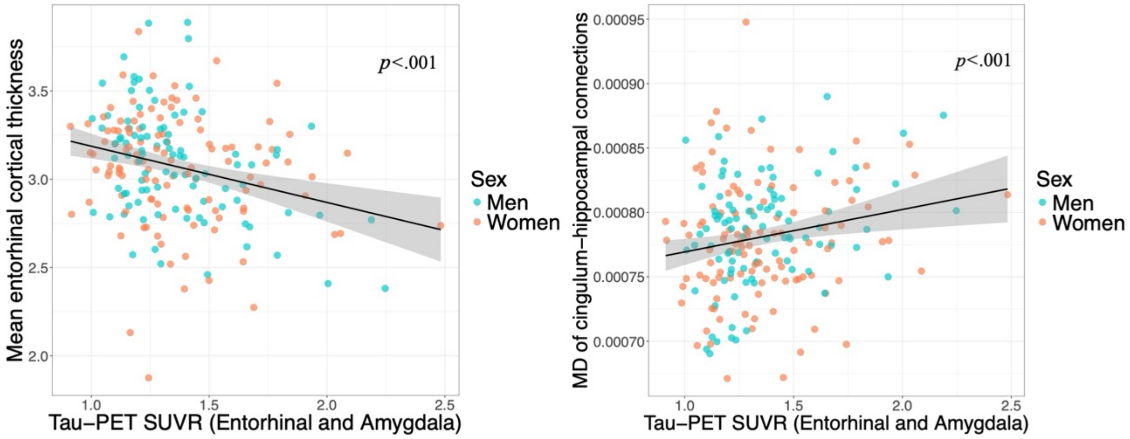


Note. Higher tau PET levels were associated with both lower entorhinal cortical thickness [β(SE)=-.375(.080), *p*<.001, $\beta_{standardized}$=-.317] and higher mean diffusivity (MD) of cingulum-hippocampal connections [β(SE)=.00005(.00001), *p*<.001, $\beta_{standardized}$=.272].

Supplemental Figure 2. Associations among men between residuals derived from full-sample models, which were then divided by sex, and those derived from sex-stratified subsamples.

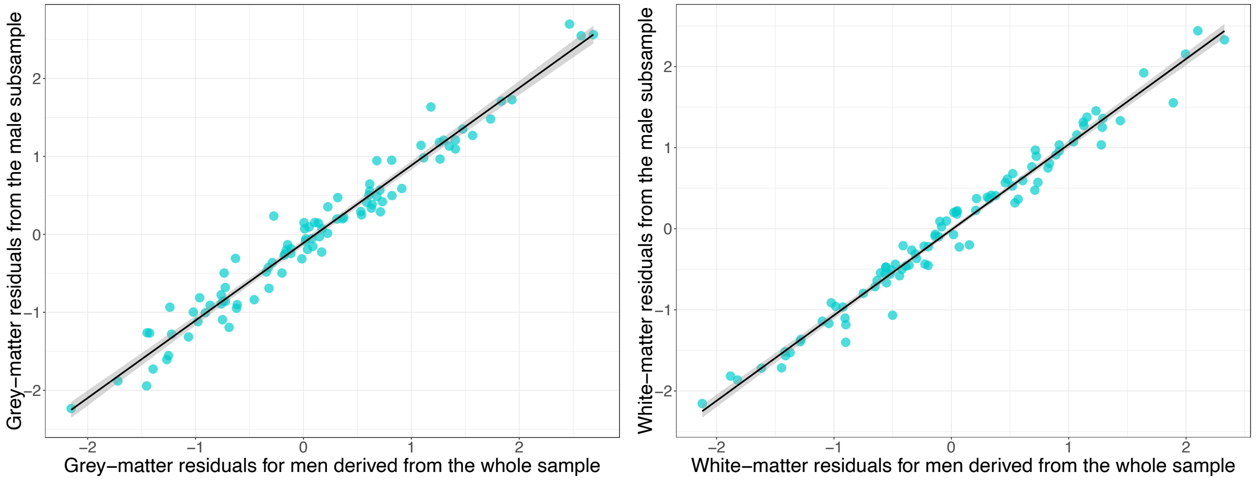


Note. The two approaches resulted in highly correlated residuals both for grey-matter resilience (*r(*91)=.98, *p*<.001) and white-matter resilience (*r(*91)=.99, *p*<.001).

Supplemental Figure 3. Associations among women between residuals derived from full-sample models, which were then divided by sex, and those derived from sex-stratified subsamples.

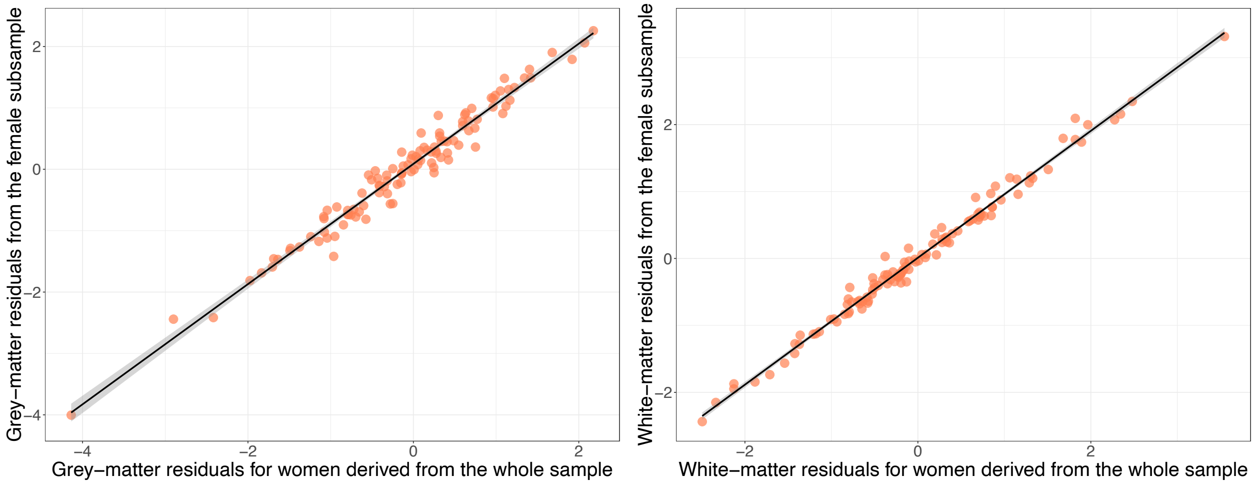


Note. The two approaches resulted in highly correlated residuals both for grey-matter resilience ($r(110)=.98$, $p<.001$) and white-matter resilience ($r(110)=.99$, $p<.001$).

Supplemental Figure 4. Association between grey- and white-matter resilience.

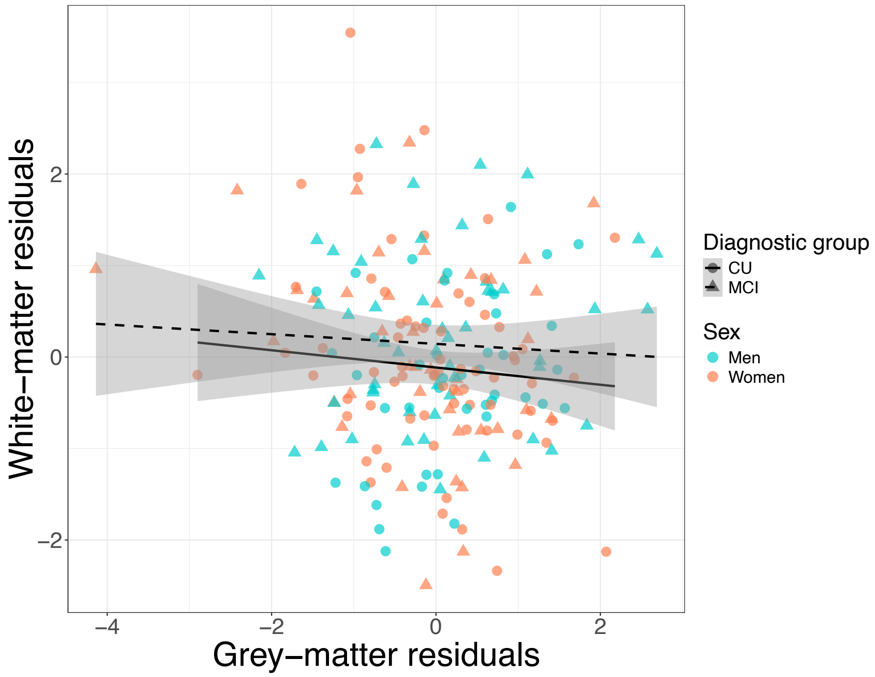


Note. There was no association between grey- and white-matter resilience ($r(203)=-.08$, $p=.273$), neither in CU ($r(111)=-.08$, $p=.373$) nor MCI individuals ($r(90)=-.06$, $p=.573$).

Supplemental Figure 5. Grey- and white-matter resilience levels by sex.

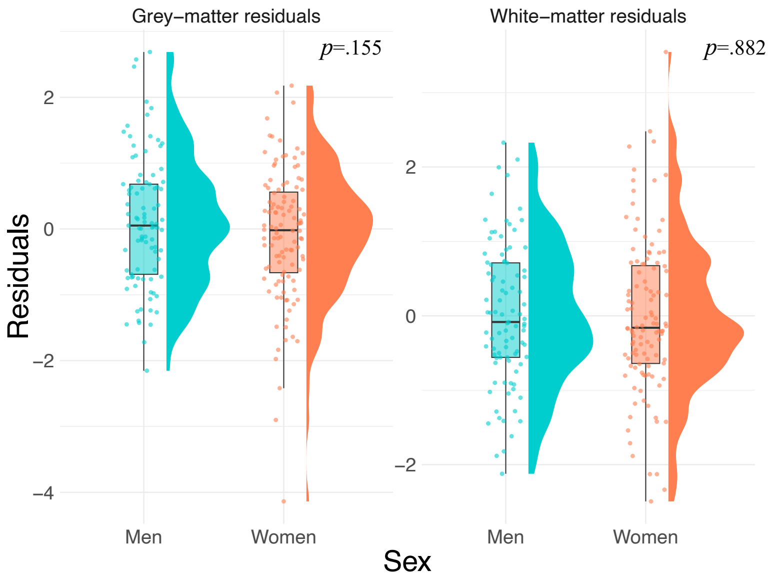


Supplemental Table 1. Associations between grey-matter resilience to tau, sex and their interaction on cognitive performance across domains.

| | Memory | Language | Executive function | Visuospatial |
|---|---|---|---|---|
| Grey-matter resilience to tau | β(SE)=.142(.047), *p*=.003**, $\beta_{standardized}$=.230 | β(SE)=.186(.054), *p*<.001***, $\beta_{standardized}$=.324 | β(SE)=.112(.058), *p*=.053, $\beta_{standardized}$=.184 | β(SE)=.105(.061), *p*=.087, $\beta_{standardized}$=.179 |
| Sex | β(SE)=.095(.066), *p*=.153, $\beta_{standardized}$=.076 | β(SE)=.009(.076), *p*=.909, $\beta_{standardized}$=.008 | β(SE)=-.060(.080), *p*=.458, $\beta_{standardized}$=-.049 | β(SE)=-.065(.086), *p*=.447, $\beta_{standardized}$=-.055 |
| Grey-matter resilience x Sex | β(SE)=-.123(.064), *p*=.054, $\beta_{standardized}$=-.148 | β(SE)=-.127(.073), *p*=.085, $\beta_{standardized}$=-.164 | β(SE)=-.172(.078), *p*=.028*, $\beta_{standardized}$=-.210 | β(SE)=-.173(.083), *p*=.038*, $\beta_{standardized}$=-.218 |
| Age | β(SE)=-.009(.005), *p*=.047*, $\beta_{standardized}$=-.104 | β(SE)=-.016(.005), *p*=.004**, $\beta_{standardized}$=-.188 | β(SE)=-.015(.006), *p*=.010*, $\beta_{standardized}$=-.168 | β(SE)=-.012(.006), *p*=.042*, $\beta_{standardized}$=-.146 |
| Education | β(SE)=.034(.013), *p*=.010**, $\beta_{standardized}$=.133 | β(SE)=.045(.015), *p*=.003**, $\beta_{standardized}$=.189 | β(SE)=.050(.016), *p*=.002**, $\beta_{standardized}$=.200 | β(SE)=.036(.017), *p*=.035*, $\beta_{standardized}$=.149 |
| Cognitive status | β(SE)=-.794(.064), *p*<.001***, $\beta_{standardized}$=-.638 | β(SE)=-.360(.074), *p*<.001***, $\beta_{standardized}$=-.312 | β(SE)=-.462(.078), *p*<.001***, $\beta_{standardized}$=-.377 | β(SE)=-.128(.083), *p*=.125, $\beta_{standardized}$=-.108 |
| $R^2_{adjusted}$ | .488 | .216 | .214 | .047 |

Note. *p<.05, **p<.01, ***p<.001. $R^2_{adjusted}$=.01, small effect size; $R^2_{adjusted}$=.09, medium effect size; $R^2_{adjusted}$ > .25, large effect size.

Supplemental Table 2. Associations between grey-matter resilience to tau and cognitive performance across domains separately in men and women.

| Men subsample | | | | |
|---|---|---|---|---|
| | Memory | Language | Executive function | Visuospatial |
| Grey-matter resilience to tau | β(SE)=.144(.049), *p*=.004**, $\beta_{standardized}$=.247 | β(SE)=.190(.051), *p*<.001***, $\beta_{standardized}$=.333 | β(SE)=.111(.059), *p*=.061, $\beta_{standardized}$=.172 | β(SE)=.120(.058), *p*=.042*, $\beta_{standardized}$=.212 |
| Age | β(SE)=-.007(.007), *p*=.365, $\beta_{standardized}$=-.077 | β(SE)=-.020(.008), *p*=.011*, $\beta_{standardized}$=-.235 | β(SE)=-.022(.009), *p*=.014*, $\beta_{standardized}$=-.226 | β(SE)=-.021(.009), *p*=.020*, $\beta_{standardized}$=-.244 |
| Education | β(SE)=.032(.019), *p*=.093, $\beta_{standardized}$=.143 | β(SE)=.047(.020), *p*=.019*, $\beta_{standardized}$=.215 | β(SE)=.071(.022), *p*=.002**, $\beta_{standardized}$=.287 | β(SE)=.030(.022), *p*=.187, $\beta_{standardized}$=.136 |
| Cognitive status | β(SE)=-.620(.096), *p*<.001***, $\beta_{standardized}$=-.536 | β(SE)=-.368(.100), *p*<.001***, $\beta_{standardized}$=-.327 | β(SE)=-.489(.114), *p*<.001***, $\beta_{standardized}$=-.383 | β(SE)=.032(.113), *p*=.779, $\beta_{standardized}$=.029 |
| $R^2_{adjusted}$ | .367 | .281 | .274 | .068 |
| **Women subsample** | | | | |
| | Memory | Language | Executive function | Visuospatial |
| Grey-matter resilience to tau | β(SE)=.010(.040), *p*=.800, $\beta_{standardized}$=.016 | β(SE)=.064(.052), *p*=.223, $\beta_{standardized}$=.109 | β(SE)=-.054(.052), *p*=.296, $\beta_{standardized}$=-.093 | β(SE)=-.066(.057), *p*=.255, $\beta_{standardized}$=-.107 |
| Age | β(SE)=-.012(.006), *p*=.050, $\beta_{standardized}$=-.128 | β(SE)=-.012(.008), *p*=.112, $\beta_{standardized}$=-.144 | β(SE)=-.011(.008), *p*=.156, $\beta_{standardized}$=-.128 | β(SE)=-.006(.008), *p*=.489, $\beta_{standardized}$=-.066 |
| Education | β(SE)=.028(.018), *p*=.120, $\beta_{standardized}$=.102 | β(SE)=.045(.023), *p*=.056, $\beta_{standardized}$=.175 | β(SE)=.032(.023), *p*=.171, $\beta_{standardized}$=.125 | β(SE)=.043(.026), *p*=.100, $\beta_{standardized}$=.158 |
| Cognitive status | β(SE)=-.955(.085), *p*<.001***, $\beta_{standardized}$=-.728 | β(SE)=-.352(.109), *p*=.002**, $\beta_{standardized}$=-.292 | β(SE)=-.453(.109), *p*<.001***, $\beta_{standardized}$=-.374 | β(SE)=-.263(.121), *p*=.032*, $\beta_{standardized}$=-.208 |
| $R^2_{adjusted}$ | .566 | .152 | .159 | .060 |

Note. *p<.05, **p<.01, ***p<.001. $R^2_{adjusted}$=.01, small effect size; $R^2_{adjusted}$=.09, medium effect size; $R^2_{adjusted}$ > .25, large effect size.

Supplemental Table 3. Associations between white-matter resilience to tau, sex and their interaction on cognitive performance across domains.

| | Memory | Language | Executive function | Visuospatial |
|---|---|---|---|---|
| White-matter resilience to tau | β(SE)=.013(.050), *p*=.803, $\beta_{standardized}$=.020 | β(SE)=.025(.059), *p*=.674, $\beta_{standardized}$=.043 | β(SE)=-.027(.062), *p*=.659, $\beta_{standardized}$=-.045 | β(SE)=.009(.065), *p*=.895, $\beta_{standardized}$=.015 |
| Sex | β(SE)=.084(.066), *p*=.204, $\beta_{standardized}$=.068 | β(SE)=-.012(.078), *p*=.879, $\beta_{standardized}$=-.010 | β(SE)=-.058(.081), *p*=.477, $\beta_{standardized}$=-.047 | β(SE)=-.062(.086), *p*=.469, $\beta_{standardized}$=-.053 |
| White-matter resilience x Sex | β(SE)=-.104(.064), *p*=.110, $\beta_{standardized}$=-.130 | β(SE)=-.061(.076), *p*=.418, $\beta_{standardized}$=-.083 | β(SE)=.032(.079), *p*=.686, $\beta_{standardized}$=.041 | β(SE)=.009(.084), *p*=.923, $\beta_{standardized}$=.011 |
| Age | β(SE)=-.008(.005), *p*=.105, $\beta_{standardized}$=-.085 | β(SE)=-.014(.005), *p*=.010*, $\beta_{standardized}$=-.172 | β(SE)=-.013(.006), *p*=.023*, $\beta_{standardized}$=-.148 | β(SE)=-.010(.006), *p*=.085, $\beta_{standardized}$=-.123 |
| Education | β(SE)=.038(.013), *p*=.004**, $\beta_{standardized}$=.150 | β(SE)=.050(.015), *p*=.001**, $\beta_{standardized}$=.210 | β(SE)=.054(.016), *p*=.001**, $\beta_{standardized}$=.214 | β(SE)=.039(.017), *p*=.024*, $\beta_{standardized}$=.160 |
| Cognitive status | β(SE)=-.794(.066), *p*<.001***, $\beta_{standardized}$=-.639 | β(SE)=-.372(.077), *p*<.001***, $\beta_{standardized}$=-.322 | β(SE)=-.452(.080), *p*<.001***, $\beta_{standardized}$=-.369 | β(SE)=-.126(.085), *p*=.142, $\beta_{standardized}$=-.106 |
| $R^2_{adjusted}$ | .478 | .166 | .195 | .026 |

Note. *p<.05, **p<.01, ***p<.001. $R^2_{adjusted}$=.01, small effect size; $R^2_{adjusted}$=.09, medium effect size; $R^2_{adjusted}$ > .25, large effect size.

Supplemental Figure 6. Associations between white-matter resilience and cognitive performance across domains separately in men (a) and women (b).

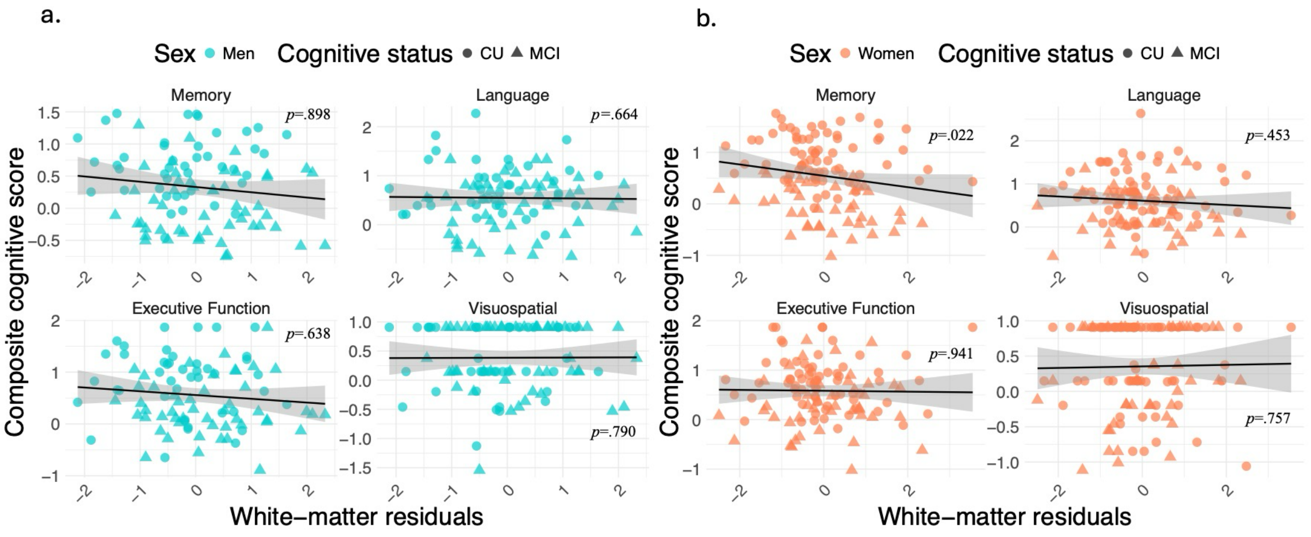

Supplemental Table 4. Associations between white-matter resilience to tau and cognitive performance across domains separately in men and women.

| Men subsample | | | | |
|---|---|---|---|---|
| | Memory | Language | Executive function | Visuospatial |
| White-matter resilience to tau | β(SE)=-.007(.055), *p*=.898, $\beta_{standardized}$=-.012 | β(SE)=.026(.059), *p*=.664, $\beta_{standardized}$=.043 | β(SE)=-.030(.064), *p*=.638, $\beta_{standardized}$=-.044 | β(SE)=-.017(.063), *p*=.790, $\beta_{standardized}$=-.029 |
| Age | β(SE)=-.004(.008), *p*=.612, $\beta_{standardized}$=-.045 | β(SE)=-.016(.008), *p*=.056, $\beta_{standardized}$=-.186 | β(SE)=-.020(.009), *p*=.027*, $\beta_{standardized}$=-.207 | β(SE)=-.018(.009), *p*=.040*, $\beta_{standardized}$=-.218 |
| Education | β(SE)=.039(.020), *p*=.050, $\beta_{standardized}$=.175 | β(SE)=.056(.021), *p*=.010*, $\beta_{standardized}$=.254 | β(SE)=.077(.023), *p*=.001**, $\beta_{standardized}$=.311 | β(SE)=.036(.023), *p*=.118, $\beta_{standardized}$=.165 |
| Cognitive status | β(SE)=-.624(.104), *p*<.001***, $\beta_{standardized}$=-.540 | β(SE)=-.390(.111), *p*<.001***, $\beta_{standardized}$=-.346 | β(SE)=-.482(.120), *p*<.001***, $\beta_{standardized}$=-.377 | β(SE)=.033(.119), *p*=.782, $\beta_{standardized}$=.030 |
| $R^2_{adjusted}$ | .306 | .172 | .246 | .024 |

| Women subsample | | | | |
|---|---|---|---|---|
| | Memory | Language | Executive function | Visuospatial |
| White-matter resilience to tau | β(SE)=-.086(.037), *p*=.022*, $\beta_{standardized}$=-.142 | β(SE)=-.037(.049), *p*=.453, $\beta_{standardized}$=-.066 | β(SE)=.004(.049), *p*=.941, $\beta_{standardized}$=.007 | β(SE)=.017(.054), *p*=.757, $\beta_{standardized}$=.029 |
| Age | β(SE)=-.011(.006), *p*=.055, $\beta_{standardized}$=-.120 | β(SE)=-.013(.008), *p*=.079, $\beta_{standardized}$=-.158 | β(SE)=-.010(.008), *p*=.208, $\beta_{standardized}$=-.113 | β(SE)=-.004(.008), *p*=.600, $\beta_{standardized}$=-.049 |
| Education | β(SE)=.030(.018), *p*=.097, $\beta_{standardized}$=.106 | β(SE)=.045(.023), *p*=.058, $\beta_{standardized}$=.175 | β(SE)=.033(.023), *p*=.166, $\beta_{standardized}$=.127 | β(SE)=.043(.026), *p*=.098, $\beta_{standardized}$=.160 |
| Cognitive status | β(SE)=-.946(.083), *p*<.001***, $\beta_{standardized}$=-.721 | β(SE)=-.360(.109), *p*=.001**, $\beta_{standardized}$=-.299 | β(SE)=-.442(.110), *p*<.001***, $\beta_{standardized}$=-.365 | β(SE)=-.252(.121), *p*=.040*, $\beta_{standardized}$=-.199 |
| $R^2_{adjusted}$ | .587 | .145 | .150 | .049 |

Note. *p<.05, **p<.01, ***p<.001. $R^2_{adjusted}$=.01, small effect size; $R^2_{adjusted}$=.09, medium effect size; $R^2_{adjusted}$ > .25, large effect size.

Supplemental Table 5. Associations between grey-matter resilience to tau, sex, and their interaction on cognitive decline over time across domains.

| | Memory | Language | Executive function | Visuospatial |
|---|---|---|---|---|
| Grey-matter resilience to tau | β(SE)=.172 (.050), *p*<.001***, $\beta_{standardized}$=.26 | β(SE)=.213 (.052), *p*<.001***, $\beta_{standardized}$=.35 | β(SE)=.127 (.057), *p*=.028*, $\beta_{standardized}$=.23 | β(SE)=.112 (.058), *p*=.056, $\beta_{standardized}$=.22 |
| Grey-matter resilience x Time | β(SE)=-.003 (.015), *p*=.868, $\beta_{standardized}$=-.006 | β(SE)=-.011 (.019), *p*=.560, $\beta_{standardized}$=-.03 | β(SE)=.006 (.020), *p*=.755, $\beta_{standardized}$=.01 | β(SE)=.010 (.024), *p*=.676, $\beta_{standardized}$=.03 |
| Sex | β(SE)=.188 (.074), *p*=.012*, $\beta_{standardized}$=.22 | β(SE)=.037 (.077), *p*=.627, $\beta_{standardized}$=.04 | β(SE)=.007 (.085), *p*=.937, $\beta_{standardized}$=.06 | β(SE)=-.054 (.086), *p*=.531, $\beta_{standardized}$=-.06 |
| Grey-matter resilience x Sex | β(SE)=-.151 (.072), *p*=.039*, $\beta_{standardized}$=-.25 | β(SE)=-.173 (.075), *p*=.022*, $\beta_{standardized}$=-.30 | β(SE)=-.178 (.083), *p*=.033*, $\beta_{standardized}$=-.22 | β(SE)=-.192 (.084), *p*=.024*, $\beta_{standardized}$=-.32 |
| Grey-matter resilience x Sex x Time | β(SE)=-.007 (.022), *p*=.758, $\beta_{standardized}$=-.01 | β(SE)=.003 (.027), *p*=.914, $\beta_{standardized}$=.007 | β(SE)=.041 (.028), *p*=.150, $\beta_{standardized}$=.10 | β(SE)=.011 (.035), *p*=.755, $\beta_{standardized}$=.03 |
| Age | β(SE)=-.005 (.005), *p*=.393, $\beta_{standardized}$=-.05 | β(SE)=-.018 (.005), *p*<.001***, $\beta_{standardized}$=-.22 | β(SE)=-.019 (.006), *p*<.001***, $\beta_{standardized}$=-.22 | β(SE)=-.012 (.005), *p*=.025*, $\beta_{standardized}$=-.15 |
| Education | β(SE)=.042 (.015), *p*=.005**, $\beta_{standardized}$=.16 | β(SE)=.041 (.014), *p*=.005**, $\beta_{standardized}$=.18 | β(SE)=.052 (.015), *p*<.001***, $\beta_{standardized}$=.22 | β(SE)=.040 (.015), *p*=.007**, $\beta_{standardized}$=.18 |
| Cognitive status | β(SE)=-.532 (.053), *p*<.001***, $\beta_{standardized}$=-.41 | β(SE)=-.311 (.059), *p*<.001***, $\beta_{standardized}$=-.27 | β(SE)=-.279 (.062), *p*<.001***, $\beta_{standardized}$=-.24 | β(SE)=-.106 (.065), *p*=.102, $\beta_{standardized}$=-.10 |

Note. *p<.05, **p<.01, ***p<.001. At baseline (time of the tau scan), the results mostly aligned with those of the cross-sectional analyses performed on the whole sample. Indeed, higher grey-matter resilience to tau was associated with better cognitive performance in memory and language, with the executive functioning domain also found significant here. Regarding the effect of sex, significant interactions in all cognitive domains also revealed that, at baseline, the protective effect of grey-matter resilience to tau on cognitive performance across domains is stronger in men. Here, women also showed better memory performance than men at baseline.

Supplemental Figure 7. Associations between grey-matter resilience and cognitive decline over time across domains separately in men (a) and women (b).

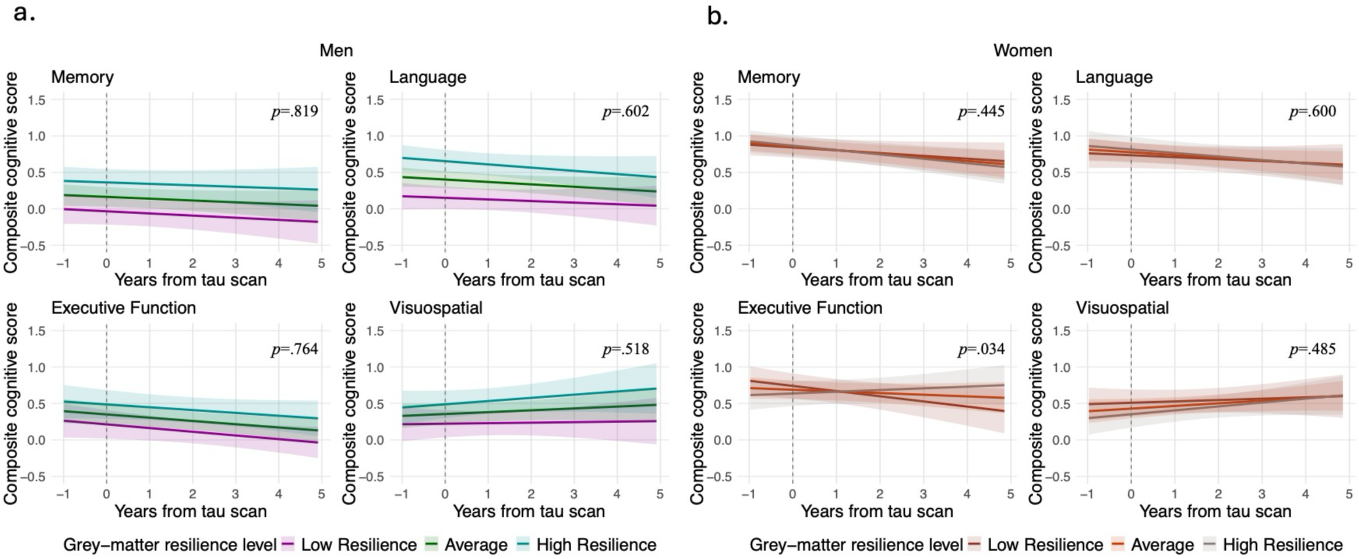


Note. The grey dashed line represents the timepoint of the tau scan. For visualization purposes only, we created a brain resilience level measure by splitting the obtained grey- and white-matter residuals into tertiles, where individuals in the bottom tertile were categorized as having "low resilience" and those in the top tertile were considered as having "high resilience".

Supplemental Table 6. Associations between grey-matter resilience to tau and cognitive decline over time across domains separately in men and women.

| Men subsample | | | | |
|---|---|---|---|---|
| | Memory | Language | Executive function | Visuospatial |
| Grey-matter resilience to tau | β(SE)=.171 (.054), *p*=.003**, $\beta_{standardized}$=.29 | β(SE)=.217 (.045), *p*<.001***, $\beta_{standardized}$=.38 | β(SE)=.118 (.059), *p*=.049*, $\beta_{standardized}$=.23 | β(SE)=.115 (.055), *p*=.042*, $\beta_{standardized}$=.25 |
| Grey-matter resilience x Time | β(SE)=.004 (.018), *p*=.819, $\beta_{standardized}$=.009 | β(SE)=-.010 (.018), *p*=.602, $\beta_{standardized}$=-.03 | β(SE)=.005 (.017), *p*=.764, $\beta_{standardized}$=.01 | β(SE)=.016 (.024), *p*=.518, $\beta_{standardized}$=.04 |
| Age | β(SE)=-.003 (.009), *p*=.752, $\beta_{standardized}$=-.03 | β(SE)=-.019 (.007), *p*=.010*, $\beta_{standardized}$=-.21 | β(SE)=-.015 (.008), *p*=.074, $\beta_{standardized}$=-.16 | β(SE)=-.017 (.008), *p*=.040*, $\beta_{standardized}$=-.19 |
| Education | β(SE)=.042 (.022), *p*=.067, $\beta_{standardized}$=.17 | β(SE)=.029 (.018), *p*=.114, $\beta_{standardized}$=.13 | β(SE)=.070 (.020), *p*<.001***, $\beta_{standardized}$=.31 | β(SE)=.040 (.020), *p*=.051, $\beta_{standardized}$=.18 |
| Cognitive status | β(SE)=-.388 (.078), *p*<.001***, $\beta_{standardized}$=-.30 | β(SE)=-.303 (.077), *p*<.001***, $\beta_{standardized}$=-.27 | β(SE)=-.273 (.081), *p*<.001***, $\beta_{standardized}$=-.23 | β(SE)=-.036 (.095), *p*=.705, $\beta_{standardized}$=-.03 |

| Women subsample | | | | |
|---|---|---|---|---|
| | Memory | Language | Executive function | Visuospatial |
| Grey-matter resilience to tau | β(SE)=.013 (.049), *p*=.796, $\beta_{standardized}$=-.002 | β(SE)=.040 (.060), *p*=.509, $\beta_{standardized}$=.04 | β(SE)=-.052 (.059), *p*=.378, $\beta_{standardized}$=.01 | β(SE)=-.079 (.063), *p*=.214, $\beta_{standardized}$=-.10 |
| Grey-matter resilience x Time | β(SE)=-.011 (.014), *p*=.445, $\beta_{standardized}$=-.02 | β(SE)=-.011 (.021), *p*=.600, $\beta_{standardized}$=-.02 | β(SE)=.048 (.022), *p*=.034*, $\beta_{standardized}$=.11 | β(SE)=.018 (.025), *p*=.485, $\beta_{standardized}$=.04 |
| Age | β(SE)=-.008 (.007), *p*=.240, $\beta_{standardized}$=-.09 | β(SE)=-.017 (.007), *p*=.023*, $\beta_{standardized}$=-.21 | β(SE)=-.020 (.008), *p*=.008**, $\beta_{standardized}$=-.24 | β(SE)=-.012 (.007), *p*=.094, $\beta_{standardized}$=-.15 |
| Education | β(SE)=.040 (.020), *p*=.050, $\beta_{standardized}$=.14 | β(SE)=.050 (.022), *p*=.027*, $\beta_{standardized}$=.20 | β(SE)=.043 (.023), *p*=.059, $\beta_{standardized}$=.17 | β(SE)=.039 (.021), *p*=.074, $\beta_{standardized}$=.17 |
| Cognitive status | β(SE)=-.659 (.072), *p*<.001***, $\beta_{standardized}$=-.51 | β(SE)=-.334 (.089), *p*<.001***, $\beta_{standardized}$=-.29 | β(SE)=-.291 (.091), *p*=.002**, $\beta_{standardized}$=-.25 | β(SE)=-.145 (.090), *p*=.110, $\beta_{standardized}$=-.13 |

Note. *p<.05, **p<.01, ***p<.001. At the time of the tau scan, the results align mostly with those of the sex-stratified cross-sectional analyses. Among men, higher grey-matter resilience is associated with better performance in all domains (executive functioning was not found significant cross-sectionally). Among women, there is no association between grey-matter resilience and cognitive performance in any domain, which also align with cross-sectional results in the female subsample.

Supplemental Table 7. Associations between white-matter resilience to tau, sex, and their interaction on cognitive decline over time across domains.

| | Memory | Language | Executive function | Visuospatial |
|---|---|---|---|---|
| White-matter resilience to tau | β(SE)=-.019 (.056), *p*=.741, $\beta_{standardized}$=-.11 | β(SE)=.021 (.060), *p*=.730, $\beta_{standardized}$=-.06 | β(SE)=-.111 (.064), *p*=.083, $\beta_{standardized}$=-.13 | β(SE)=.052 (.066), *p*=.434, $\beta_{standardized}$=.07 |
| White-matter resilience x Time | β(SE)=-.043 (.015), *p*=.006**, $\beta_{standardized}$=-.09 | β(SE)=-.045 (.019), *p*=.022*, $\beta_{standardized}$=-.11 | β(SE)=.027 (.021), *p*=.208, $\beta_{standardized}$=.06 | β(SE)=-.009 (.026), *p*=.728, $\beta_{standardized}$=-.02 |
| Sex | β(SE)=.171 (.076), *p*=.026*, $\beta_{standardized}$=.20 | β(SE)=.014 (.080), *p*=.858, $\beta_{standardized}$=.02 | β(SE)=.009 (.085), *p*=.920, $\beta_{standardized}$=.04 | β(SE)=-.055 (.087), *p*=.526, $\beta_{standardized}$=-.06 |
| White-matter resilience x Sex | β(SE)=-.091 (.074), *p*=.222, $\beta_{standardized}$=-.06 | β(SE)=-.077 (.079), *p*=.331, $\beta_{standardized}$=-.07 | β(SE)=.059 (.084), *p*=.479, $\beta_{standardized}$=.02 | β(SE)=-.077 (.086), *p*=.371, $\beta_{standardized}$=-.11 |
| White-matter resilience x Sex x Time | β(SE)=.044 (.020), *p*=.034*, $\beta_{standardized}$=.09 | β(SE)=.030 (.026), *p*=.248, $\beta_{standardized}$=.07 | β(SE)=-.039 (.028), *p*=.168, $\beta_{standardized}$=-.09 | β(SE)=.011 (.035), *p*=.754, $\beta_{standardized}$=.03 |
| Age | β(SE)=-.003 (.005), *p*=.526, $\beta_{standardized}$=-.04 | β(SE)=-.017 (.005), *p*=.002**, $\beta_{standardized}$=-.20 | β(SE)=-.018 (.006), *p*=.001**, $\beta_{standardized}$=-.21 | β(SE)=-.010 (.005), *p*=.053, $\beta_{standardized}$=-.13 |
| Education | β(SE)=.048 (.015), *p*=.002**, $\beta_{standardized}$=.18 | β(SE)=.049 (.015), *p*=.002**, $\beta_{standardized}$=.21 | β(SE)=.059 (.015), *p*<.001***, $\beta_{standardized}$=.25 | β(SE)=.044 (.015), *p*=.003**, $\beta_{standardized}$=.19 |
| Cognitive status | β(SE)=-.528 (.053), *p*<.001***, $\beta_{standardized}$=-.41 | β(SE)=-.313 (.061), *p*<.001***, $\beta_{standardized}$=-.27 | β(SE)=-.276 (.062), *p*<.001***, $\beta_{standardized}$=-.24 | β(SE)=-.120 (.067), *p*=.073, $\beta_{standardized}$=-.11 |

Note. *p<.05, **p<.01, ***p<.001. At the time of the tau scan (baseline), the results aligned with those of the cross-sectional analyses performed on the whole sample. Indeed, white-matter resilience to tau was not associated with cognitive performance in any domain. Regarding the effect of sex, non-significant interactions in all cognitive domains also revealed that, at baseline, the association between white-matter resilience to tau and cognitive performance did not vary by sex. Here, women also showed better memory performance than men at baseline.

Supplemental Table 8. Associations between white-matter resilience to tau and cognitive decline over time across domains separately in men and women.

| Men subsample | | | | |
|---|---|---|---|---|
| | Memory | Language | Executive function | Visuospatial |
| White-matter resilience to tau | β(SE)=-.039 (.064), *p*=.541, $\beta_{standardized}$=-.13 | β(SE)=.020 (.058), *p*=.729, $\beta_{standardized}$=-.06 | β(SE)=-.116 (.066), *p*=.086, $\beta_{standardized}$=-.13 | β(SE)=.049 (.065), *p*=.457, $\beta_{standardized}$=.06 |
| White-matter resilience x Time | β(SE)=-.038 (.017), *p*=.027*, $\beta_{standardized}$=-.08 | β(SE)=-.045 (.018), *p*=.014*, $\beta_{standardized}$=-.11 | β(SE)=.029 (.018), *p*=.114, $\beta_{standardized}$=.07 | β(SE)=-.009 (.027), *p*=.739, $\beta_{standardized}$=-.02 |
| Age | β(SE)=.0002 (.009), *p*=.981, $\beta_{standardized}$=.002 | β(SE)=-.015 (.008), *p*=.073, $\beta_{standardized}$=-.17 | β(SE)=-.013 (.008), *p*=.134, $\beta_{standardized}$=-.14 | β(SE)=-.014 (.008), *p*=.099, $\beta_{standardized}$=-.15 |
| Education | β(SE)=.053 (.024), *p*=.029*, $\beta_{standardized}$=.22 | β(SE)=.043 (.021), *p*=.044*, $\beta_{standardized}$=.20 | β(SE)=.080 (.021), *p*<.001***, $\beta_{standardized}$=.36 | β(SE)=.049 (.021), *p*=.022*, $\beta_{standardized}$=.22 |
| Cognitive status | β(SE)=-.388 (.079), *p*<.001***, $\beta_{standardized}$=-.30 | β(SE)=-.304 (.084), *p*<.001***, $\beta_{standardized}$=-.27 | β(SE)=-.264 (.082), *p*=.002**, $\beta_{standardized}$=-.23 | β(SE)=-.078 (.100), *p*=.440, $\beta_{standardized}$=-.07 |
| Women subsample | | | | |
| | Memory | Language | Executive function | Visuospatial |
| White-matter resilience to tau | β(SE)=-.109 (.042), *p*=.012*, $\beta_{standardized}$=-.18 | β(SE)=-.056 (.054), *p*=.303, $\beta_{standardized}$=-.13 | β(SE)=-.052 (.053), *p*=.330, $\beta_{standardized}$=-.12 | β(SE)=-.026 (.058), *p*=.657, $\beta_{standardized}$=-.05 |
| White-matter resilience x Time | β(SE)=.0008 (.013), *p*=.949, $\beta_{standardized}$=.002 | β(SE)=-.014 (.018), *p*=.434, $\beta_{standardized}$=-.04 | β(SE)=-.011 (.021), *p*=.589, $\beta_{standardized}$=-.03 | β(SE)=.0008 (.023), *p*=.970, $\beta_{standardized}$=.002 |
| Age | β(SE)=-.008 (.006), *p*=.188, $\beta_{standardized}$=-.09 | β(SE)=-.018 (.007), *p*=.017*, $\beta_{standardized}$=-.21 | β(SE)=-.021 (.007), *p*=.005, $\beta_{standardized}$=-.25 | β(SE)=-.011 (.007), *p*=.119, $\beta_{standardized}$=-.14 |
| Education | β(SE)=.040 (.019), *p*=.044*, $\beta_{standardized}$=.14 | β(SE)=.050 (.022), *p*=.025*, $\beta_{standardized}$=.20 | β(SE)=.043 (.022), *p*=.059, $\beta_{standardized}$=.17 | β(SE)=.038 (.021), *p*=.079, $\beta_{standardized}$=.16 |
| Cognitive status | β(SE)=-.668 (.071), *p*<.001***, $\beta_{standardized}$=-.52 | β(SE)=-.344 (.088), *p*<.001***, $\beta_{standardized}$=-.30 | β(SE)=-.303 (.091), *p*=.001**, $\beta_{standardized}$=-.26 | β(SE)=-.139 (.090), *p*=.125, $\beta_{standardized}$=-.13 |

Note. *p<.05, **p<.01, ***p<.001. At the time of the tau scan, the results align with those of the cross-sectional analyses in the male subsample, such that white-matter resilience is not associated with cognitive performance in any domain. Among women, there is an association between higher white-matter resilience to tau and better memory performance at baseline, which also align with cross-sectional results in the female subsample.
